# Association of air pollution from a landfill site with primary care consultation

**DOI:** 10.1101/2024.09.16.24313470

**Authors:** Kelvin P Jordan, Sara Muller, Muhammad Usman, James Bailey, Claire Burton, Sarah A Lawton, Christian D Mallen, Kayleigh J Mason, Simon Wathall, Danielle A van der Windt

## Abstract

**Background:** Waste landfill sites are associated with gaseous emissions and this air pollution can cause unpleasant smells (“malodour”). This causes concerns about its impact on the health of the local population. This study assessed change in general practice consultation behaviour during a period of increased complaints associated with air pollution at a UK landfill site.

**Methods:** The study period was October 2020 to December 2021. The age-sex standardised prevalence and incidence of consultations for mental health, respiratory, and other symptoms hypothesised to be impacted by the air pollution issues were determined and compared between: i) 6 practices located close to the landfill site (zone A), ii) 6 practices located a mid-distance from the site (zone B), iii) 6 practices located further away and expected to have had less impact (zone C).

**Results:** Whilst there was an increased consultation for mental health problems in practices nearest to the landfill site compared to those furthest away, consultation frequencies for respiratory and other potentially associated symptoms were lower and likelihood of consultation was consistently highest in practices located in zone B.

**Conclusion:** This study did not show clear evidence of an increase in recorded primary healthcare contacts for conditions and symptoms hypothesised to be connected to air pollution. It highlighted the challenges of examining the impact of air pollution on the health of local populations. Since this study focussed on coded consultations in primary care and not symptoms present in the general population, an impact on the health of individuals cannot be ruled out.

**How this fits in:**

- Air pollution from waste landfill sites may impact on the health of the local population.
- We did not find consistent evidence of increased healthcare consultations to general practices which were nearest to such pollution.
- This study highlights the challenges in examining the impact of air pollution on health.
- Symptoms may still be increased in the general population and an impact on the health of individuals cannot be ruled out.

## Introduction

Waste management is a challenging undertaking, requiring consideration of the consequences to human health and well-being, environmental preservation, sustainability, and economy.(1) Hazardous waste includes substances such as asbestos, chemicals, solvents, oils, and hazardous waste containers. Guidance exists to ensure that waste is identified, classified, and coded before sent for further treatment (if hazardous), recycling or disposal.(2)

The characterisation including type and origin, composition, leachability and other properties determines its disposal destination. In the UK, each landfill site has agreed waste acceptance criteria (WAC). Any hazardous waste that must be landfilled and cannot meet WAC is ‘problematic waste’ and requires communication with the Environment Agency.(3) The criteria and procedures for the acceptance and ongoing management and monitoring of waste at landfills is highly regulated.(4,5) Whilst such regulations exist, the World Health Organisation has raised concerns about the possible health impacts of waste management.(1)

Landfill sites are associated with gaseous emissions which arise from physical, chemical, and biological processes occurring within the waste (e.g. microbial production, chemical reaction and volatilisation). ‘Landfill gas’ includes methane, carbon dioxide, nitrogen, oxygen, hydrogen, and trace components, the composition and density of which will depend on the waste and nature of the site. Gases emitted from landfill sites in high enough concentrations can cause toxicity, corrosion, ecotoxity, and malodour. Emission levels are therefore monitored and regulated, and gas management systems deployed to protect human health and the environment.(6) Malodorous emissions can be an annoyance to local communities and a cause of complaints due to public concern about their impact on health and quality of life.(6)

For several years there have been reports of malodour allegedly linked to a landfill site in England, increasing from October 2020 and peaking between January and September 2021. This has resulted in a high level of complaints to the Environment Agency and local council. Given concerns about environmental pollution, its potential detrimental impact on health, and anecdotal reports of increased utilisation of healthcare for related symptoms, this study aimed to determine the association of this air pollution with changes in local primary care consultation patterns for potentially relevant symptoms and health conditions.

## Methods

### Study setting

The study comprised of three zones of participating general practices.

Zone A (exposed practices) consisted of six practices selected from within the immediate vicinity (3km) of the landfill site. Patients consulting at these practices were hypothesised to live closest to the site and hence more likely to have experienced an impact on their health.

Zone B (near practices) consisted of six practices between 3km and 8km from the site. These were selected to match their combined registered population as much as possible to zone A by neighbourhood deprivation and ethnicity.

Zone C (control practices) consisted of six practices selected from those located between 8km and 40km from the alleged source, and not close to another landfill site. These were also selected to match their combined registered population as much as possible to zone A by neighbourhood deprivation and ethnicity. Patients from these practices were expected to have had less impact from the site.

For the purposes of identifying suitable near and control practices, deprivation and ethnicity data was extracted from the Office for Health Improvement and Disparities National General Practice Profiles.(7) Deprivation was based on the 2019 Index of Multiple Deprivation, and ethnicity based on the weighted average of non-white ethnic groups from the 2011 Census data over the contributing lower-level Super Output Areas (LSOAs). These give a general representation of the local population.

### Study population

The study population was patients (all ages) consulting between 1 January 2017 and 31 December 2021 for one of the outcome conditions at the general practices. Patients were excluded if they had asked for their medical records not to be used for research and had a relevant code indicating this in their records.

### Outcomes

The study period was October 2020 (when complaints to the Environment Agency started to escalate) to December 2021 with the baseline period set at January 2017-September 2020. Outcomes were coded consultations at the general practices for three categories of “case” conditions hypothesised by consensus of study team and stakeholders to have greater levels of consultation if health had been impacted by air pollution. These conditions were:

i. Mental health (depression, anxiety, stress, panic attacks, insomnia, hypersomnia, somnolence, peri- and post-natal mental health conditions);
ii. Respiratory (asthma, chronic obstructive pulmonary disease, breathlessness, wheeze, cough);
iii. Other symptoms (nausea, headache/migraine, eye inflammation/irritation, sore throat, rhinitis/sinusitis, hay fever, dizziness, epistaxis). Two “control” conditions hypothesised not to be related to any air pollution/environmental issues were investigated to allow assessment of whether any increase in consultation for the case conditions is due to an increased propensity to consult. These conditions were:
iv. Urinary tract infection (UTI);
v. Epilepsy.

The conditions were defined by SNOMED concept IDs, used in UK general practice to record morbidities and symptoms for which patients present, with code lists modified from other studies and consensus of the study team. The code lists are given in the supplementary files.

Anonymised consultations with a relevant SNOMED concept ID for one of the case or control conditions were extracted from each practice, including patient year of birth and sex. A count of the registered population by age (ten-year intervals) and sex, at the start of 2020 was obtained for each participating practice to utilise as the denominator population.

### Analysis

Data analysis was performed for i) all people consulting for a condition (prevalence) and ii) restricted to those consulting with a new episode (incidence) defined as no consultation within the same category (e.g., mental health; respiratory) in the previous 12 months. Consultation rates were standardised to the age-sex population structure of the local region.

Within each group of practices monthly age-sex standardised prevalence and incidence per 1,000 registered population for each case and control condition were determined. A graphical comparison of trends over time was performed with focus on the study period being from October 2020. Joinpoint regression was used to examine changes in trends in consultation rates. Joinpoint regression identifies months where a statistically significant change (the “joinpoint”) in the underlying monthly percentage change (MPC) in consultation rates occurred. The most appropriate number of joinpoints was based on the lowest weighted Bayesian information criterion (BIC). It was expected there would be a joinpoint showing a decreasing trend in consultation around the start of the COVID-19 pandemic in March 2020 with a second joinpoint in the summer of 2020 indicating recovery of consultation rates. We hypothesised that a bigger MPC would be seen in zone A following the summer 2020 joinpoint if there was an association with air pollution.

Difference in differences analyses were performed to evaluate whether monthly consultation prevalence and incidence after October 2020 varied between zones, adjusting for baseline (pre-October 2020) rates. A multilevel (month within practice) negative binomial model was used for each study outcome. Time was categorised i) dichotomously i.e., pre-October 2020 (baseline) and post-October 2020 and ii) into four periods i.e., pre-October 2020 (baseline), October-December 2020, January-September 2021, and October-December 2021. January-September 2021 was the peak period for odour complaints. Prevalence and incidence were modelled as a function of zones, time, and an interaction term of zones with time. Risk Ratios (RRs) of relative differences between the consultation rates in zones over time along with 95% confidence intervals (CI) are reported. Finally, we graphically compared monthly volume of complaints recorded by the Environment Agency with consultation prevalence.

### Sample size

Taking mental health as the example, assuming a total population of 42,000 across the 6 practices in each zone, and monthly consultation prevalence of 8 per 1,000 registered population, this will give a 95% CI of ±1/1,000 for the monthly consultation prevalence.

Approval was obtained from the University Research Ethics Committee (REC Project Reference 0138) and Health Research Authority (HRA) (Reference: 22/PR/0078).

## Results

All invited practices agreed to take part. Registered practice populations for the zones were 46,178 (zone A), 42,678 (zone B) and 45,882 (zone C). More of the registered population were aged 60 and over in zone C (zone A: 25%; zone B: 28%; zone C: 31%). Zone C had a lower non-white ethnic population (zone A: 6%; zone B: 4%; zone C: 2%) whilst the majority of practices across the three zones served populations on average in mid-deprived neighbourhoods with zone A having a slightly higher population from more deprived neighbourhoods.

Figure 1 shows monthly prevalence. For all conditions there is a fall in prevalence around the start of the COVID-19 pandemic (March 2020). For mental health, zone B has the highest prevalences both before and after October 2020 with zone A fluctuating between higher and similar rates to zone C. Zone B also has the highest prevalences of respiratory and other symptoms following October 2020. For the control conditions there is more variation in trend over time post-October 2020 in the monthly consultation prevalence and no consistent differences between zones can be seen. There were smaller absolute differences between zones in monthly incidence (Supplementary Figure 1).

**Figure 1.**
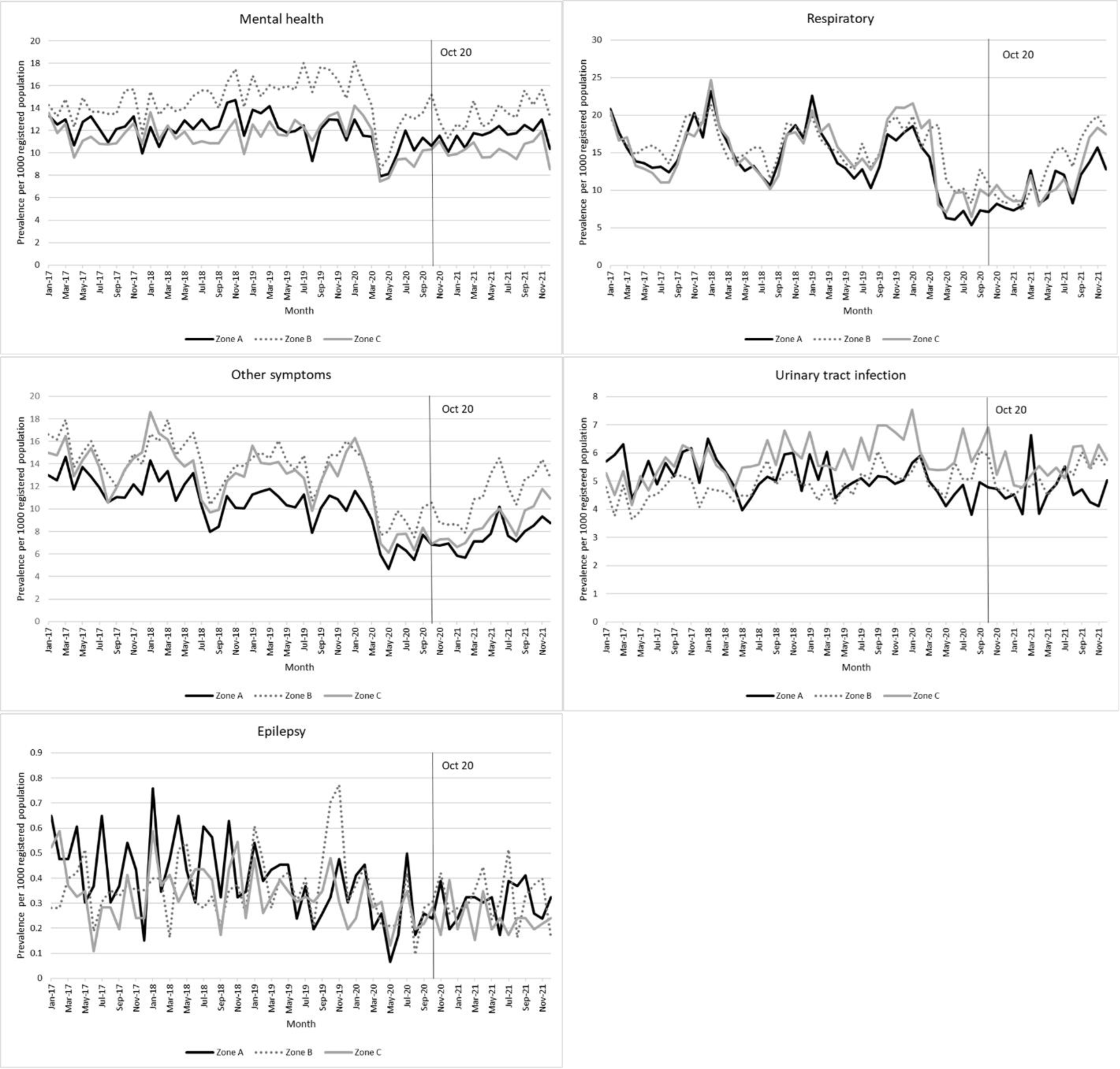
Age-sex standardised monthly consultation prevalence for each health condition.

There was a joinpoint at the time of the first COVID-19 lockdown indicating a substantial drop in prevalence of consultations for respiratory and other symptoms with an upturn in consultations starting from May-December 2020. From October 2020, MPC was higher for zone B practices for respiratory (A 4.98% increase per month, B 7.94%, C 3.48%) and similar across zones for other symptoms (2.15-2.73%) (Table 1). For incidence, MPC was similar for zone A and B for mental health (A 1.20%, B 1.39%, C 0.52%), highest for zone B for respiratory (A 7.57%, B 9.01%, C 8.95%) and similar across zones for other symptoms (2.30-2.92%).

**Table 1.** Date of last joinpoint post March 2020 and subsequent monthly percentage change in consultation rates.

|  | Zone A | Zone B | Zone C |
| --- | --- | --- | --- |
| Prevalence |  |  |  |
| Mental health | <sup>a</sup> | <sup>a</sup> | <sup>a</sup> |
| Respiratory | June 2020: 4.98% | December 2020: 7.94% | May 2020: 3.48% |
| Other symptoms | May 2020: 2.15% | May 2020: 2.73% | May 2020: 2.57% |
| Urinary tract infection | <sup>a</sup> | <sup>a</sup> | <sup>a</sup> |
| Epilepsy | <sup>a</sup> | <sup>a</sup> | <sup>a</sup> |
| Incidence |  |  |  |
| Mental health | April 2020: 1.20% | April 2020: 1.39% | April 2020: 0.52% |
| Respiratory | July 2020: 7.57% | July 2020: 9.01% | August 2020: 8.95% |
| Other symptoms | May 2020: 2.30% | May 2020: 2.92% | May 2020: 2.89% |
| Urinary tract infection | <sup>a</sup> | <sup>a</sup> | <sup>a</sup> |
| Epilepsy | <sup>a</sup> | <sup>a</sup> | <sup>a</sup> |

<sup>a</sup> No joinpoint detected

Difference in differences analyses of monthly prevalence with dichotomous categorisation of study period (pre- and post-October 2020) are presented in Table 2. Monthly prevalence of mental health consultations was higher in zone A relative to zone C, adjusting for pre-October 2020 rates (RR 1.13; 95% CI 1.09, 1.17) post-October 2020, but it was lower for zone A for respiratory (RR 0.90; 95% CI 0.83, 0.97) and other symptoms (RR 0.87; 95% CI 0.84, 0.91). Zone A had higher prevalence of epilepsy than zone C but lower prevalence of UTI.

**Table 2.** Associations of zone with consultation prevalence in the period October 2020-December 2021.

|  | **Mean monthly prevalence**<br>**per 1,000** |  |  | **Zone A vs Zone B**<br>**(Zone B is reference)** |  |  | **Zone A vs Zone C**<br>**(Zone C is reference)** |  |  | **Zone B vs Zone C**<br>**(Zone C is reference)** |  |  |
| --- | --- | --- | --- | --- | --- | --- | --- | --- | --- | --- | --- | --- |
|  | **Zone A** | **Zone B** | **Zone C** | **RR** | **Lower**<br>**95% CI** | **Upper**<br>**95% CI** | **RR** | **Lower**<br>**95% CI** | **Upper**<br>**95% CI** | **RR** | **Lower**<br>**95% CI** | **Upper**<br>**95% CI** |
| Mental health | 11.53 | 13.56 | 10.23 | 0.85 | 0.82 | 0.88 | 1.13 | 1.09 | 1.17 | 1.33 | 1.27 | 1.38 |
| Respiratory | 10.34 | 13.01 | 11.49 | 0.79 | 0.73 | 0.86 | 0.90 | 0.83 | 0.97 | 1.13 | 1.04 | 1.24 |
| Other symptoms | 7.57 | 11.28 | 8.68 | 0.67 | 0.65 | 0.70 | 0.87 | 0.84 | 0.91 | 1.30 | 1.25 | 1.35 |
| Urinary tract infection | 4.69 | 5.17 | 5.62 | 0.91 | 0.83 | 0.99 | 0.83 | 0.76 | 0.92 | 0.92 | 0.88 | 0.96 |
| Epilepsy | 0.35 | 0.39 | 0.28 | 0.88 | 0.77 | 1.00 | 1.23 | 1.03 | 1.47 | 1.40 | 1.18 | 1.67 |

Age-sex standardised. RR = risk ratio adjusted for consultations rates January 2017-September 2020.

For all three case conditions (mental health, respiratory, and other symptoms) zone A had lower prevalence rates compared to zone B post-October 2020 (mental health zone A vs B: RR 0.85, 95% CI 0.82, 0.88; respiratory 0.79, 95% CI 0.73, 0.86; other symptoms 0.67; 0.65, 0.70). Among control conditions zone A had slightly lower prevalence rates of UTI compared to zone B, whereas there was no difference for epilepsy post-October 2020.

Prevalences were higher in zone B compared to zone C post-October 2020 for all case conditions. Patterns for the case conditions were similar when focusing on the January– September 2021 period when complaints were highest except there was no difference in consultation for respiratory conditions in zones A and B versus zone C (Supplementary Table 1).

Difference in differences analyses of monthly incidence with dichotomous categorisation of study period is presented in Table 3. Zone A had a higher incidence of mental health conditions than zone C post-October 2020 but a lower incidence of respiratory symptoms with no difference for other symptoms and the control conditions. Incidence of respiratory and other symptoms post-October 2020 were higher in zone B compared to zone A, but there was no difference in consultations for mental health conditions. There was greater monthly consultation for epilepsy in zone A relative to zone B. Zone B practices had higher incidence post-October 2020 than zone C for all three case conditions but lower for UTI. Estimates were similar when focussing on the January-September 2021 period although the increased incidence for zone A versus zone C for mental health conditions was not statistically significant (Supplementary Table 2).

**Table 3.** Associations of zone with consultation incidence in the period October 2020-December 2021.

|  | Mean monthly incidence<br>per 1,000 |  |  | Zone A vs Zone B<br>(Zone B is reference) |  |  | Zone A vs Zone C<br>(Zone C is reference) |  |  | Zone B vs Zone C<br>(Zone C is reference) |  |  |
| --- | --- | --- | --- | --- | --- | --- | --- | --- | --- | --- | --- | --- |
|  | Zone A | Zone B | Zone C | RR | Lower<br>95% CI | Upper<br>95% CI | RR | Lower<br>95% CI | Upper<br>95% CI | RR | Lower<br>95% CI | Upper<br>95% CI |
| Mental health | 2.22 | 2.23 | 2.03 | 1.00 | 0.92 | 1.08 | 1.10 | 1.01 | 1.19 | 1.10 | 1.00 | 1.21 |
| Respiratory | 1.64 | 2.48 | 2.13 | 0.66 | 0.61 | 0.73 | 0.77 | 0.73 | 0.82 | 1.17 | 1.08 | 1.25 |
| Other symptoms | 2.32 | 2.78 | 2.35 | 0.84 | 0.77 | 0.91 | 0.99 | 0.92 | 1.06 | 1.18 | 1.09 | 1.28 |
| Urinary tract infection | 1.60 | 1.53 | 1.73 | 1.05 | 0.95 | 1.15 | 0.93 | 0.83 | 1.04 | 0.89 | 0.80 | 0.99 |
| Epilepsy | <sup>a</sup> | <sup>a</sup> | <sup>a</sup> | 1.65 | 1.19 | 2.29 | 1.15 | 0.75 | 1.78 | 0.70 | 0.41 | 1.18 |

Age-sex standardised. RR=risk ratio adjusted for consultations rates January 2017-September 2020 <sup>a</sup> Not reported due to small numbers

Plots of monthly consultation prevalence (Figure 2) against volume of complaints showed no indication of higher consultation in months with a higher level of complaints. This was also the case for incidence (Supplementary Figure 2).

**Figure 2.**
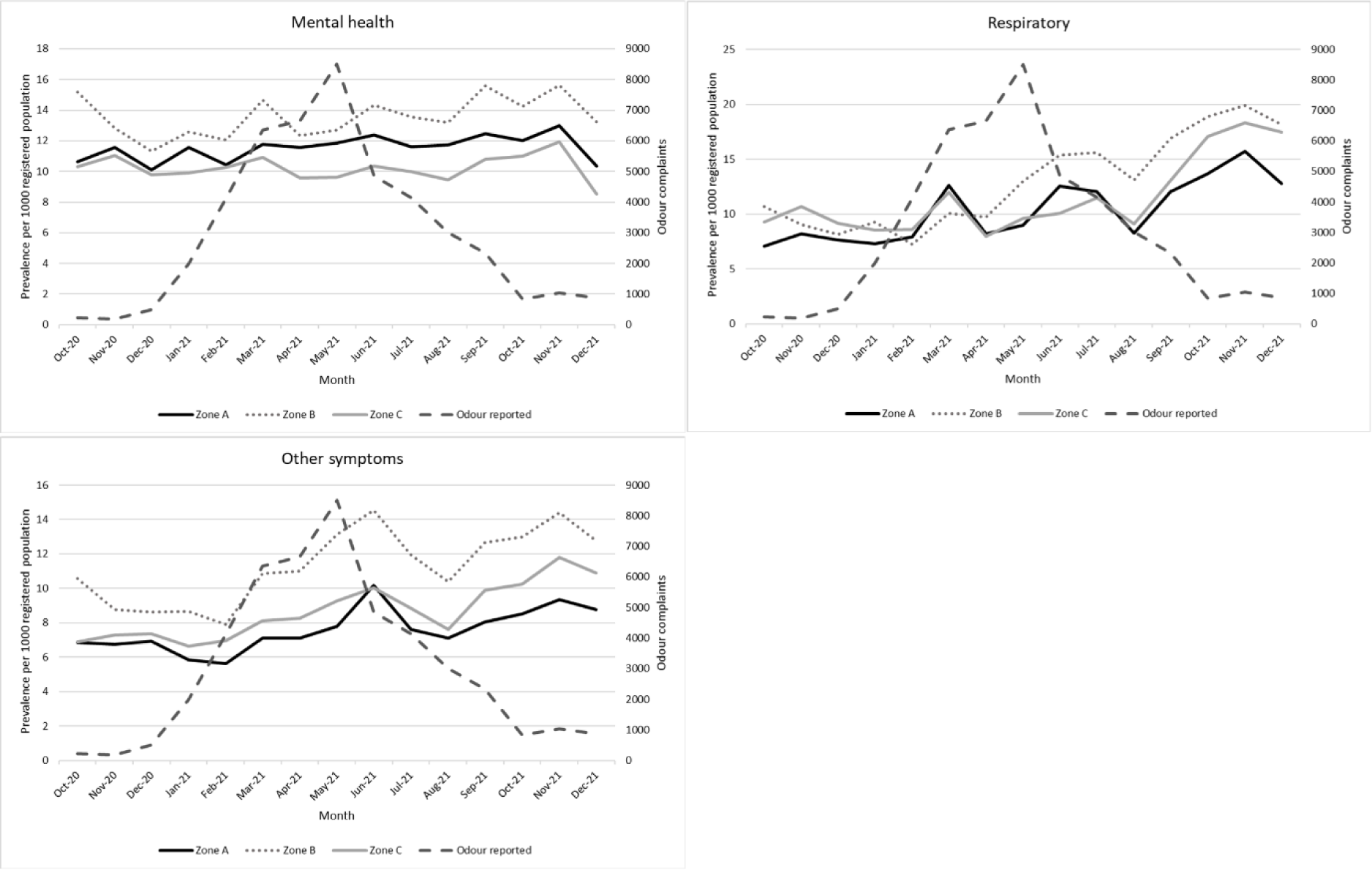
Age-sex standardised monthly consultation prevalence vs volume of odour complaints post-October 2020.

## Discussion

This study examined whether patients registered at general practices closest to a landfill site with high volumes of complaints of malodour consulted more frequently for symptoms that might be expected to be associated with air pollution. Whilst patients at practices closest to the site consulted more for mental health problems than patients at practices furthest away, the highest consultation rates generally occurred for patients at practices located a mid-distance away.

Systematic reviews have found some evidence of an association with poorer health of living near hazardous waste, including respiratory and mental health symptoms.(8-12) Similarly, there is some evidence of an association of air pollution with mental health problems.(13) However, these reviews have generally concluded evidence is limited with quality of studies needing to be improved. For example, many studies examining the impact of waste management on health suffer from limitations due to poor exposure assessment, ecological level of analysis, or lack of information on relevant confounders including areas near waste sites may already have health inequalities.(8,11,14) One review found some evidence of an increased risk of mortality, respiratory diseases, and negative mental health effects associated with residing near landfills.(11) However, in many cases, the evidence was inadequate to establish a strong relationship with the outcomes. Another review concluded that living close to a well-managed landfill site should pose no significant health risks, however highlighted the annoyance resulting from any malodour from landfill sites may cause mental and physical symptoms.(14)

There was little evidence in this study to suggest an increase in consultation for relevant symptoms close to the landfill site. However, the study and prior reviews highlight the challenges of assessing the impact of air pollution at landfill sites. We focussed on one such site where complaints have been high, and the methods and findings can inform studies assessing impact at other sites across the UK. The limitations indicated below though means that we cannot infer that there has been no impact on the health of the local population. In particular, it is likely that some people had relevant symptoms but chose not to seek health care. This study also only measured health care use coded in primary care and individuals may have sought other forms of care, for example, emergency services. The longer-term impact on health needs to be evaluated.

Strengths of this study were the large denominator population and all the practices invited consented to being part of the study. We used an objective measure of health, namely recorded reasons for consultation at the general practices. We examined two control conditions which we do not expect to be associated with air pollution and the malodour. We adjusted for baseline differences between practices.

A limitation was that the COVID-19 pandemic had a substantial impact on consultation behaviour.(15) Patient concerns about access to primary care and risks of exposure to COVID-19 may have influenced the decision to consult, particularly at the start of the outcome period (October 2020). The symptoms of COVID-19 also overlap with symptoms reported to be associated with the malodour. However, we have no reason to believe consultation behaviours or COVID-19 related presentations would be different across the three zones. We did not assess severity of symptoms. We could not examine the free text which gives more information about the consultation and may indicate whether the patient or clinician linked the symptom to the landfill site. Proximity to site was based on location of the practices and some patients may live some distance from their registered practice. We did not take account of the direction of residence from the landfill site, which may influence the likelihood of odour exposure. The symptoms groups we used were broad and may not have captured differences for specific symptoms between zones. This study can only assess strength of association of air pollution with consultation and not causation.

## Conclusion

Whilst there was no clear evidence of an increase in consultation for symptoms connected to air pollution near a landfill site, the impact on the health of the local population cannot be ruled out, particularly given that this study focussed on coded primary care consultations and did not investigate symptoms in the general population. Further research should investigate the longer-term impact on health including self-reported symptoms.

## Supporting information

Supplementary codelists

## Acknowledgements

The general practices who agreed to take part, the NHS Commissioning Support Unit (CSU), who undertook the searches at the participating practices, and Alec Dobney (UK Health Security Agency, formally Public Health England) for advice on study design.

## Ethics

University Research Ethics Committee REC Project Reference 0138. Health Research Authority (HRA) approvals (Reference: 22/PR/0078).

## Data Availability

The datasets underlying this article cannot be shared publicly due to privacy of individuals as they refer to individual consultation records. However summarised data are available from the corresponding author on reasonable request.

