## Supplementary tables and figures for "Association of air pollution from a landfill site with primary care consultation"

**Supplementary Figure 1 – Age-sex standardised monthly consultation incidence for each health condition**

**(Epilepsy not presented due to low numbers)**

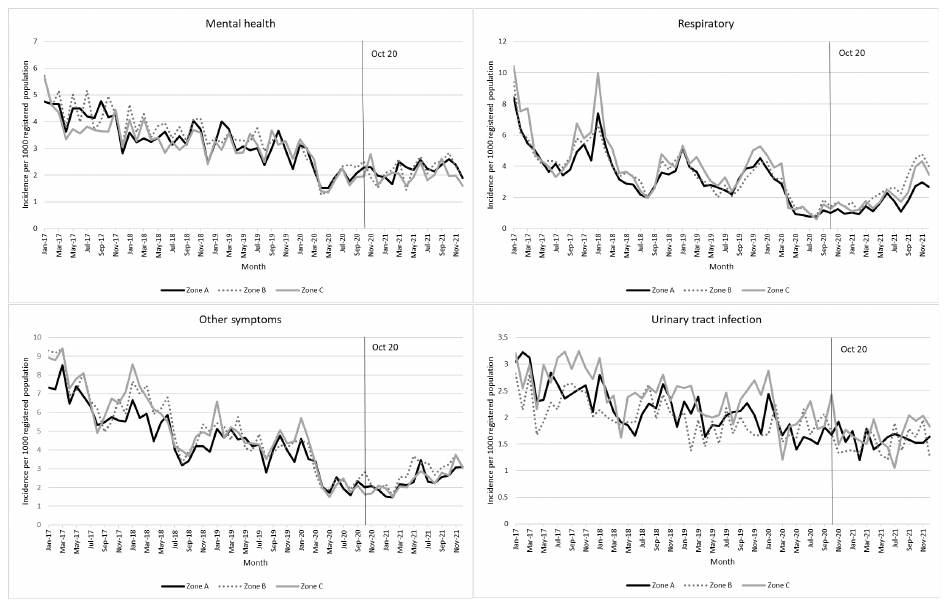

**Supplementary Figure 2 – Age-sex standardised monthly consultation incidence vs volume of odour complaints post-October 2020**

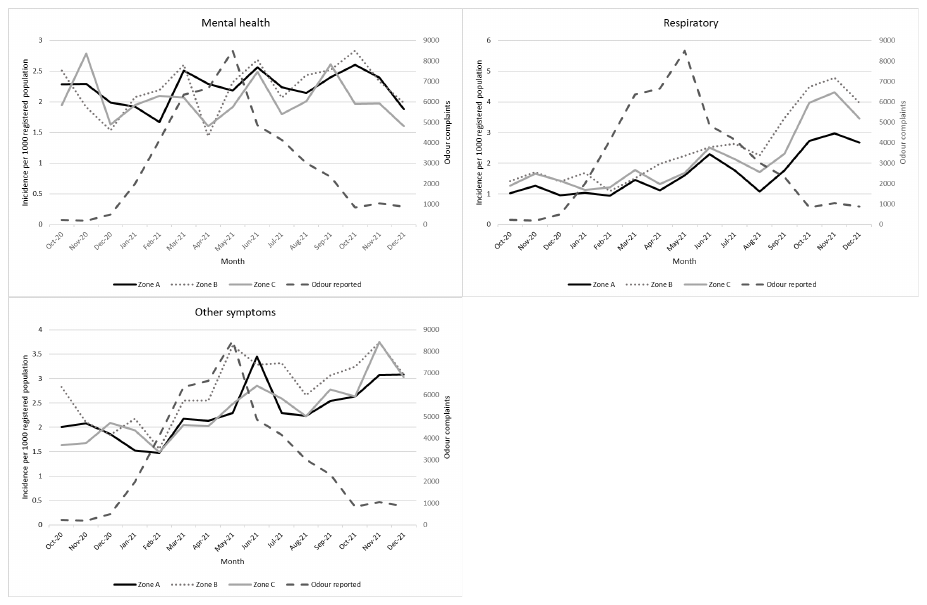

**Supplementary Table 1 Associations of zone with consultation prevalence between October-December 2021, split into three periods**

|  | **Zone A vs Zone B**  **(Zone B is reference)** | | | **Zone A vs Zone C**  **(Zone C is reference)** | | | **Zone B vs Zone C**  **(Zone C is reference)** | | |
| --- | --- | --- | --- | --- | --- | --- | --- | --- | --- |
|  | **RR** | **Lower 95% CI** | **Upper 95% CI** | **RR** | **Lower 95% CI** | **Upper 95% CI** | **RR** | **Lower 95% CI** | **Upper 95% CI** |
| October-December 2020 | | | | | | | | | |
| Mental health | 0.82 | 0.71 | 0.94 | 1.04 | 1.03 | 1.05 | 1.26 | 1.11 | 1.44 |
| Respiratory | 0.82 | 0.69 | 0.98 | 0.79 | 0.75 | 0.82 | 0.96 | 0.82 | 1.12 |
| Other symptoms | 0.74 | 0.66 | 0.82 | 0.95 | 0.92 | 0.98 | 1.30 | 1.13 | 1.49 |
| Urinary tract infection | 0.91 | 0.81 | 1.01 | 0.76 | 0.67 | 0.87 | 0.84 | 0.80 | 0.89 |
| Epilepsy | 0.83 | 0.78 | 0.89 | 0.93 | 0.65 | 1.34 | 1.12 | 0.82 | 1.52 |
| January-September 2021 | | | | | | | | | |
| Mental health | 0.87 | 0.84 | 0.90 | 1.16 | 1.11 | 1.21 | 1.33 | 1.28 | 1.38 |
| Respiratory | 0.82 | 0.72 | 0.93 | 0.99 | 0.93 | 1.07 | 0.96 | 0.82 | 1.12 |
| Other symptoms | 0.66 | 0.63 | 0.68 | 0.88 | 0.84 | 0.92 | 1.34 | 1.28 | 1.39 |
| Urinary tract infection | 0.95 | 0.84 | 1.07 | 0.89 | 0.78 | 1.01 | 0.93 | 0.89 | 0.98 |
| Epilepsy | 0.93 | 0.80 | 1.08 | 1.37 | 1.10 | 1.71 | 1.48 | 1.21 | 1.81 |
| October-December 2021 | | | | | | | | | |
| Mental health | 0.82 | 0.79 | 0.85 | 1.12 | 1.06 | 1.18 | 1.37 | 1.25 | 1.49 |
| Respiratory | 0.74 | 0.70 | 0.78 | 0.80 | 0.74 | 0.86 | 1.08 | 1.05 | 1.11 |
| Other symptoms | 0.66 | 0.65 | 0.68 | 0.81 | 0.79 | 0.83 | 1.22 | 1.18 | 1.26 |
| Urinary tract infection | 0.80 | 0.70 | 0.91 | 0.77 | 0.67 | 0.88 | 0.96 | 0.94 | 0.98 |
| Epilepsy | 0.78 | 0.52 | 1.17 | 1.20 | 1.08 | 1.34 | 1.55 | 0.98 | 2.45 |

Age-sex standardised. RR=risk ratio adjusted for consultations rates January 2017-September 2020

**Supplementary Table 2 Associations of zone with consultation incidence between October-December 2021, split into three periods**

|  | **Zone A vs Zone B**  **(Zone B is reference)** | | | **Zone A vs Zone C**  **(Zone C is reference)** | | | **Zone B vs Zone C**  **(Zone C is reference)** | | |
| --- | --- | --- | --- | --- | --- | --- | --- | --- | --- |
|  | **RR** | **Lower 95% CI** | **Upper 95% CI** | **RR** | **Lower 95% CI** | **Upper 95% CI** | **RR** | **Lower 95% CI** | **Upper 95% CI** |
| October-December 2020 | | | | | | | | | |
| Mental health | 1.10 | 0.92 | 1.32 | 1.03 | 0.83 | 1.28 | 0.94 | 0.68 | 1.29 |
| Respiratory | 0.71 | 0.69 | 0.74 | 0.74 | 0.68 | 0.81 | 1.04 | 0.99 | 1.09 |
| Other symptoms | 0.88 | 0.73 | 1.07 | 1.10 | 0.92 | 1.32 | 1.25 | 0.91 | 1.72 |
| Urinary tract infection | 1.17 | 0.97 | 1.41 | 0.91 | 0.67 | 1.22 | 0.78 | 0.69 | 0.87 |
| Epilepsy | 1.06 | 0.52 | 2.15 | 0.58 | 0.24 | 1.43 | 0.55 | 0.14 | 2.11 |
| January-September 2021 | | | | | | | | | |
| Mental health | 0.98 | 0.88 | 1.09 | 1.07 | 0.97 | 1.19 | 1.10 | 1.03 | 1.17 |
| Respiratory | 0.67 | 0.57 | 0.79 | 0.83 | 0.77 | 0.89 | 1.23 | 1.08 | 1.39 |
| Other symptoms | 0.81 | 0.72 | 0.91 | 0.98 | 0.91 | 1.07 | 1.21 | 1.14 | 1.30 |
| Urinary tract infection | 1.03 | 0.93 | 1.15 | 0.98 | 0.84 | 1.14 | 0.95 | 0.82 | 1.11 |
| Epilepsy | 1.72 | 1.24 | 2.38 | 1.60 | 1.02 | 2.51 | 0.93 | 0.53 | 1.65 |
| October-December 2021 | | | | | | | | | |
| Mental health | 0.96 | 0.91 | 1.02 | 1.24 | 1.17 | 1.31 | 1.29 | 1.16 | 1.43 |
| Respiratory | 0.63 | 0.60 | 0.66 | 0.71 | 0.67 | 0.76 | 1.13 | 1.11 | 1.15 |
| Other symptoms | 0.87 | 0.79 | 0.97 | 0.93 | 0.83 | 1.05 | 1.07 | 0.96 | 1.18 |
| Urinary tract infection | 0.97 | 0.76 | 1.24 | 0.81 | 0.74 | 0.88 | 0.83 | 0.71 | 0.97 |
| Epilepsy | 2.48 | 1.14 | 5.43 | 1.04 | 0.42 | 2.61 | 0.42 | 0.28 | 0.64 |

Age-sex standardised. RR=risk ratio adjusted for consultations rates January 2017-September 2020
